# Efficacy and safety of treating symptomatic, primary focal osteochondral lesions of the patella with a combination of intra-articular injection of hyaluronic acid (HAi) and radial extracorporeal shock wave therapy (rESWT) or HAi alone: a retrospective two-cohort study

**DOI:** 10.1101/2020.07.29.20164111

**Authors:** Jin Cao, Heqin Huang, Changgui Zhang, Liu Yang, Christoph Schmitz, Xiaojun Duan

## Abstract

**Background:** This study tested the hypothesis that a combination of intra-articular injection of hyaluronic acid (HAi) and radial extracorporeal shock wave therapy (rESWT) is more effective than HAi alone in the treatment of symptomatic, primary focal osteochondral lesions of the patella (sPFOLP).

**Methods:** We performed a retrospective analysis of n=81 subjects with unilateral sPFOLP who were treated with respectively HAi+rESWT (n=36) or HAi alone (n=45) at the outpatient department of our hospital between January 1^st^, 2014 and January 31^th^, 2018. Subjects in both cohorts also received education with special attention to reduction of going up and down the stairs, squatting and specific activities, as well as non-opioid analgesics if necessary. Efficacy was assessed using the pain visual analogue scale (VAS) and the Western Ontario and McMaster Universities Osteoarthritis Index (WOMAC) score, which were examined at baseline (BL) and six weeks (W6), three months (M3) and six months (M6) after BL, as well as during a last follow-up examination (LE) which was performed at 37.6 ± 1.7 (mean ± standard error of the mean) months after BL (range, 12-59 months). In addition, measurements of the area of the patellar bone marrow edema (PBME) on sagittal, fat suppression sequence magnetic resonance imaging (MRI) scans of the knee were performed at BL, M3, M6 and LE.

**Results:** Subjects who were treated with HAi+rESWT showed significantly smaller mean VAS pain scores at W6, M3 and M6, significantly smaller mean WOMAC scores at W6, M3 and LE and significantly smaller mean PBME areas at M3, M6 and LE than subjects who were treated with HAi alone. No severe adverse events were observed.

**Discussion:** The results of this study suggest that HAi+rESWT is safe and more effective than HAi alone in the treatment of sPFOLP. This result should be verified in adequate randomized controlled trials.

**Level of evidence:** Level III; retrospective two-cohort study.

## BACKGROUND

Osteochondral lesions of the knee, which are often affiliated with trauma or degeneration, are very common [1–5]. The patella has been identified as the most common location of chondromalacia [6]. Focal chondral and osteochondral lesions of the patella are characterized by knee dysfunction and pain, which is aggravated when going up and down the stairs and squatting [5]. The disease may substantially limit common movements in daily life, such as rising from a chair, going up and down the stairs and climbing hills.

The pathogenesis of osteochondral lesions of the patella is still under investigation. It has been hypothesized that adverse stress stimulation or local microcirculation disturbance may be critically involved in the pathogenesis of symptomatic, primary focal osteochondral lesions of the patella (sPFOLP) [7].

The main complaint in the early stage of sPFOLP is pain in the absence of obvious abnormalities observed on radiographs, whereas magnetic resonance imaging (MRI) scans can show focal avascular necrosis or signals of bone marrow edema (BME) in the subchondral bone of the patella [7, 8]. With progression of sPFOLP radiographs and MRI scans can show gradually narrowing of the patellofemoral joint space, cartilage surface defects, subchondral bone sclerosis and cystic degeneration [8]. In case of severe degeneration the final progress may be osteoarthritis (OA) of the whole knee joint.

Current treatment options of sPFOLP focus on reducing certain activities in daily life including going up and down the stairs, squatting, strenuous running and jumping, as well as local or oral use of analgesics, physical therapy, etc. [9]. Another option is intra-articular injection of hyaluronic acid (HAi) or platelet rich plasma into the knee joint [9–11]. However, these treatments do often not result in long-lasting pain relief and improvement of function [9, 10]. Surgical treatment options include bone marrow stimulation, implantation of autologous chondrocytes, minced cartilage scaffolds, osteochondral autografts, osteochondral allografts or structural synthetic scaffolds, and patellofemoral arthroplasty [9, 12]. However, these treatments have some drawbacks, including the potential for complications, a lengthy recovery and a failure rate of up to 23% in case of osteochondral allograft transplantation [9, 13]. Thus, there is need to identify and validate novel therapeutic options for improving treatment efficacy and slowing down the progression of sPFOLP.

Over the past decades extracorporeal shock wave therapy (ESWT) has emerged as a promising treatment for many musculoskeletal conditions [14–16]. In particular, ESWT has shown therapeutic value for treating avascular necrosis of the femoral head [17–20] and osteochondral lesions of the talus [21].

We started to treat sPFOLP with a combination of HAi and radial extracorporeal shock wave therapy (rESWT) at our hospital (Center for Joint Surgery, Southwest Hospital, Third Military Medical University (Army Medical University), Chongqing, China) already in 2014. Subjects who do not want to be treated with rESWT are offered HAi alone.

This retrospective study evaluated the efficacy and safety of treating sPFOLP with respectively HAi+rESWT or HAi alone. The hypotheses were that (i) treatment of sPFOLP with HAi+rESWT is more effective than HAi alone, and (ii) treatment of sPFOLP with HAi+rESWT does not result in any serious adverse event..

## METHODS

### Study design

This was a retrospective study comparing HAi+rESWT with HAi alone for the management of sPFOLP. All subjects were treated at the outpatient department of the Center for Joint Surgery, Southwest Hospital, Third Military Medical University (Army Medical University), Chongqing, China between January 1^st^, 2014 and January 31^st^, 2018.

### Ethics

This retrospective study was approved by the Ethics Committee of the Southwest Hospital, Third Military Medical University (Army Medical University), Chongqing, China (No. KY201915). All procedures that involved human participants were performed in accordance with the ethical standards of the institutional and national research committee and with the 1975 Helsinki declaration and its later amendments or comparable ethical standards. Written informed consent was obtained from all individual participants included in this study.

### Participants

All female and male adults aged 18-65 years with unilateral sPFOLP who were treated with respectively HAi+rESWT or HAi alone at the outpatient department of the Center for Joint Surgery, Southwest Hospital, Third Military Medical University (Army Medical University), Chongqing, China between January 1st, 2014 and January 31st, 2018 were eligible for being enrolled in this study. There were no subjects in the hospital’s database who had suffered from sPFOLP and were treated with rESWT alone.

Diagnosis of sPFOLP was based on the following criteria: unilateral or bilateral pain of the knee joint for more than three months, particularly when going up and down the stairs and relieved after rest; aggravation of symptoms over time; no history of perceived local trauma; presence of a focal patellar bone marrow edema and/or localized osteonecrosis evidenced by MRI; and exclusion of rheumatoid arthritis, gout, tuberculosis and other inflammatory diseases.

Exclusion criteria for being enrolled in this study were: bilateral sPFOLP (because in case of bilateral sPFOLP pain perception could be different from pain perception in case of unilateral sPFOLP), joint degeneration with radiographs showing narrow joint space / Kellgren-Lawrence Grades II to IV, valgus or varus deformity >5 degrees of the knee, previous treatments with intra-articular injections and/or ESWT (both radial and focused), contraindications of rESWT (signs of local infection and/or tumor, serious blood dyscrasia, blood-clotting disorders, treatment with oral anticoagulants) and incomplete availability of radiographs and MRI images of the affected knee joint in the hospital’s database for confirmation of diagnosis and evaluation of treatment outcome.

### Interventions

All subjects diagnosed with sPFOLP were provided with a detailed description of the benefits and potential risks of treatment selection prior to treatment. All subjects were offered HAi+rESWT or HAi alone; no subject was offered rESWT alone. In select cases subjects were offered surgery (not considered in this study). In addition, all subjects received education with special attention to reduction of going up and down the stairs, squatting and specific activities, as well as non-opioid analgesics (Celecoxib 200 mg, oral, once per day) if necessary.

We identified n=36 subjects in the hospital’s database with unilateral sPFOLP who had made an informed decision for being treated with HAi+rESWT. These subjects received five intra-articular injections of 2.5 mL HA (ARTZ Dispo, Seikagaku Corporation, Tokyo, Japan) each into the affected knee joint, with one injection per week. In addition, subjects received rESWT immediately before each HA injection, which was performed with the Swiss DolorClast device (EMS Electro Medical Systems, Nyon, Switzerland) using the EvoBlue handpiece and the 15-mm applicator. Each rESWT session consisted of 2000 radial extracorporeal shock waves (rESWs) applied to the area of the patellar osteochondral lesion as determined by MRI in sagittal and axial positions. Tender points around the patella served as auxiliary treatment areas. The knee flexion was 45 degrees during rESWT. The positive energy flux density (ESW^+^) of the rESWs was between 0.04 and 0.08 mJ/mm^2^, corresponding to an air pressure of 1.8-2.5 bar. The rESWs were applied at a frequency of 6-8 rESWs/second. No anesthesia or analgesic drugs were applied during the rESWT sessions. All rESWT sessions were performed by the same therapist.

Furthermore, we identified n=45 subjects in the hospital’s database with unilateral sPFOLP who had made an informed decision for being treated with HAi alone. Injection of HA was performed as described for the subjects who were treated with HAi+rESWT.

Baseline characteristics of the subjects are summarized in Table 1.

**Table 1.**
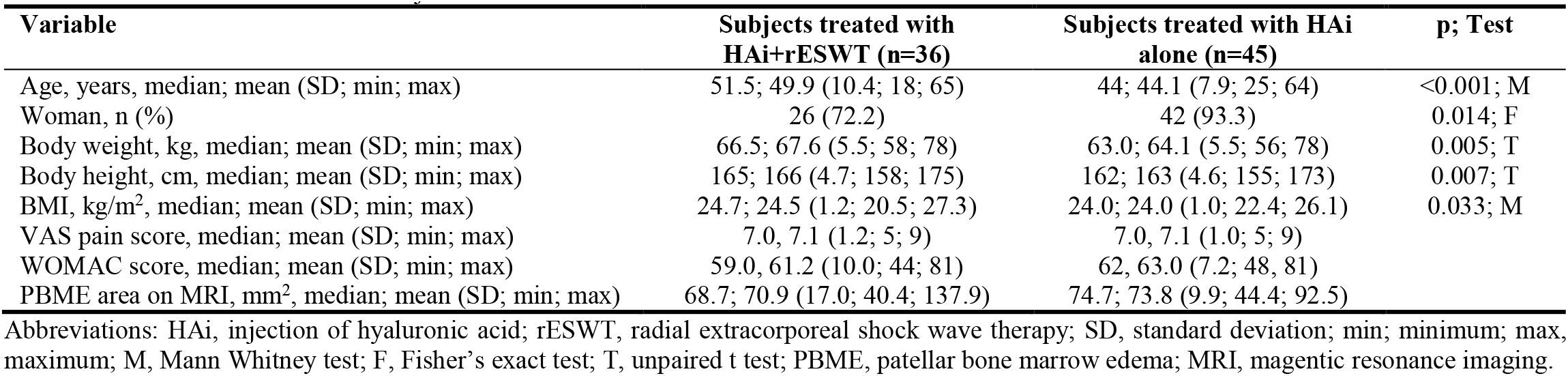
Characteristics of included subjects at baseline.

### Outcome measurements and assessments

The primary clinical outcome was pain during normal daily life activities, measured on a visual analogue scale (VAS) ranging from zero (no pain at all) to ten (maximum, unbearable pain). The VAS pain score was assessed by the same examiner at baseline (BL) and six weeks (W6), three months (M3) and six months (M6) after BL, as well as during a last follow-up examination (LE) that took place at 37.6 ± 1.7 (mean ± standard error of the mean; SEM) months after BL (range, 12-59 months).

Treatment success was defined as individual improvement of the VAS pain score by at least 60% at M3.

Secondary clinical outcomes were the Western Ontario and McMaster University Osteoarthritis Index (WOMAC) score and measurements of the area of the patellar bone marrow edema (PBME) on magnetic resonance imaging (MRI) scans of the knee. The WOMAC score was assessed by the same examiner at BL, W6, M3, M6 and LE. Magnetic resonance imaging was performed with an 0.2 Tesla Artoscan-C device (Esaote, Genoa, Italy) at BL, M3, M6 and LE. The Materialise Mimics software (Version 19.0; Materialise, Leuven, Belgium) was used to determine the maximum PBME area on sagittal, fat suppression sequence images by two independent, experienced radiologists, whose results were averaged.

### Statistical analysis

For all investigated variables, mean and SEM were calculated at BL, W6, M3, M6 and LE. This was separately performed for the subjects who were treated with HAi+rESWT and the subjects who were treated with HAi alone. Because this was a retrospective two-cohort study, tests of baseline differences between the two cohorts were performed (according to [22] tests of baseline differences in randomized controlled trials (RCTs) are illogical). After determining using the D’Agostino and Pearson omnibus normality test whether the distribution of the variables age, body weight, body height and body mass index (BMI) of the subjects in both cohorts at BL were consistent with a Gaussian distribution, baseline comparisons were performed with unpaired Student’s t test (mean body weight and mean body height), Mann Whitney test (mean age and mean BMI) and Fisher’s exact test (gender). These tests showed statistically significant differences between the two cohorts at BL (Table 1).

Accordingly, changes in mean VAS pain score, mean WOMAC score and mean PBME area over time after treatment were assessed using the general linear model repeated measures (GLM-RM) procedure, with treatment as fixed factor, the investigated variable (VAS pain score, WOMAC score, PBME area) as dependent variable, and the subjects’ gender, age, body weight, body height and BMI as covariates. Then, the post hoc Mann Whitney test was applied for pairwise comparisons. Because each data set was used in two statistical analyses (GLM-RM procedure and post-hoc test), in all analyses an effect was considered statistically significant if its associated P value was smaller than 0.025. Calculations were performed using SPSS (Version 26; IBM, Armonk, NY, USA) and GraphPad Prism (Version 8.2.1; GraphPad software, San Diego, CA, USA).

Treatment success (i.e., number of subjects with individual improvement in VAS pain score by at least 60%) was tested with Chi-square test.

No subject was lost during the follow-up period, i.e., all subjects passed the examinations at BL, W6, M3, M6 and LE. As a result, complete capture of all data from all subjects was achieved.

## RESULTS

### VAS pain score

The mean VAS pain score of the subjects who were treated with HAi+rESWT decreased from 7.1 ± 0.2 at BL to 3.8 ± 0.1 at W6, 3.0 ± 0.1 at M3, 2.4 ± 0.1 at M6 and 1.6 ± 0.1 at LE, and of the subjects who were treated with HAi alone from 7.1 ± 0.1 (mean ± SEM) at BL to 5.2 ± 0.1 at W6, 3.9 ± 0.1 at M3, 2.9 ± 0.1 at M6 and 2.0 ± 1.2 at LE (Fig. 1A). The GLM-RM procedure showed a statistically significant interaction between Time and Treatment (Table 2). The Mann Whitney test showed statistically significant differences between the cohorts at W6, M3 and M6 (Table 2).

**Table 2.** Results of statistical analysis (p values).

|  | VAS Pain Score | WOMAC Score | PBME Area |
| --- | --- | --- | --- |
| **Results of General Linear Model Repeated Measures procedure** |  |  |  |
| Time | 0.327 | 0.811 | 0.414 |
| Time × Gender | **0.011** | 0.079 | 0.110 |
| Time × Age | 0.753 | 0.815 | 0.485 |
| Time × Height | 0.327 | 0.815 | 0.374 |
| Time × Weight | 0.255 | 0.780 | 0.443 |
| Time × BMI | 0.282 | 0.802 | 0.530 |
| Time × Treatment | **< 0.001** | **< 0.001** | **< 0.001** |
| **Results of post-hoc pairwise comparisons** |  |  |  |
| BL | 0.653 | 0.398 | 0.048 |
| W6 | **< 0.001** | **< 0.001** |  |
| M3 | **< 0.001** | **< 0.001** | **< 0.001** |
| M6 | **0.005** | 0.112 | **< 0.001** |
| LE | 0.038 | **0.006** | **< 0.001** |

Abbreviations: PBME, patellar bone marrow edema; BL, baseline; W6, six weeks after baseline; M3, three months after baseline; M6, six months after baseline; LE, time point of the last follow-up examination (between twelve and 59 months after baseline). P values < 0.025 are given in boldface.

**Figure 1.**
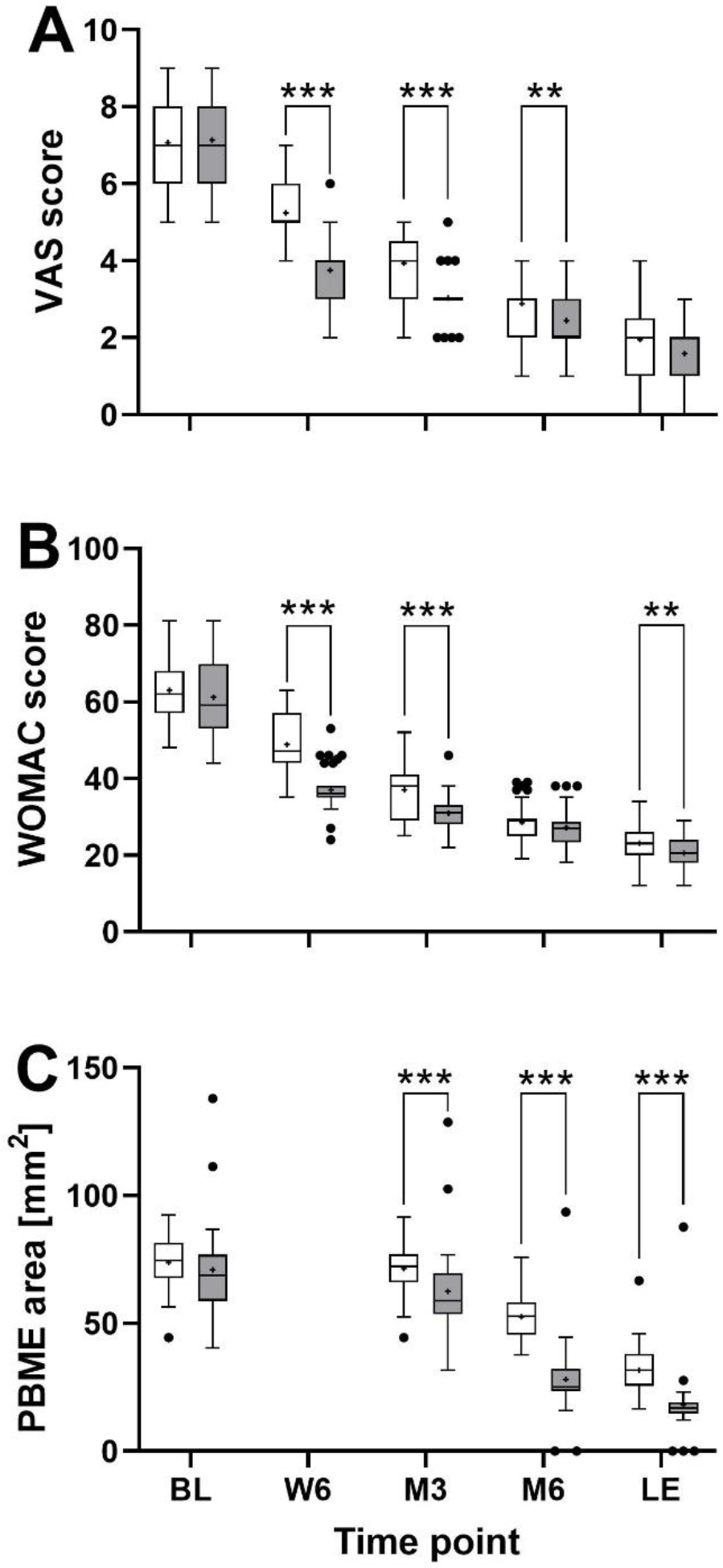
Tukey boxplots of VAS pain score (A), WOMAC score (B) and the area of patella bone marrow edema (PBME) detected on sagittal, fat suppression sequence MRI scans (C) of subjects with unilateral, symptomatic, primary focal osteochondral lesions of the patella who were treated with respectively a combination of injection of hyaluronic acid (HAi) and radial extracorporeal shock wave therapy (gray bars) or HAi alone (open bars). Data were collected at baseline (BL), six weeks (W6), three months (M3) and six months (M6) after baseline, as well as at a last follow-up examination (LE) which was performed at 37.6 ± 1.7 (mean ± standard error of the mean) months after BL (range, 12-59 months). Results of pairwise comparisons using the Mann Whithney test are indicated (**, p < 0.01; ***, p < 0.001).

**Figure 2.**
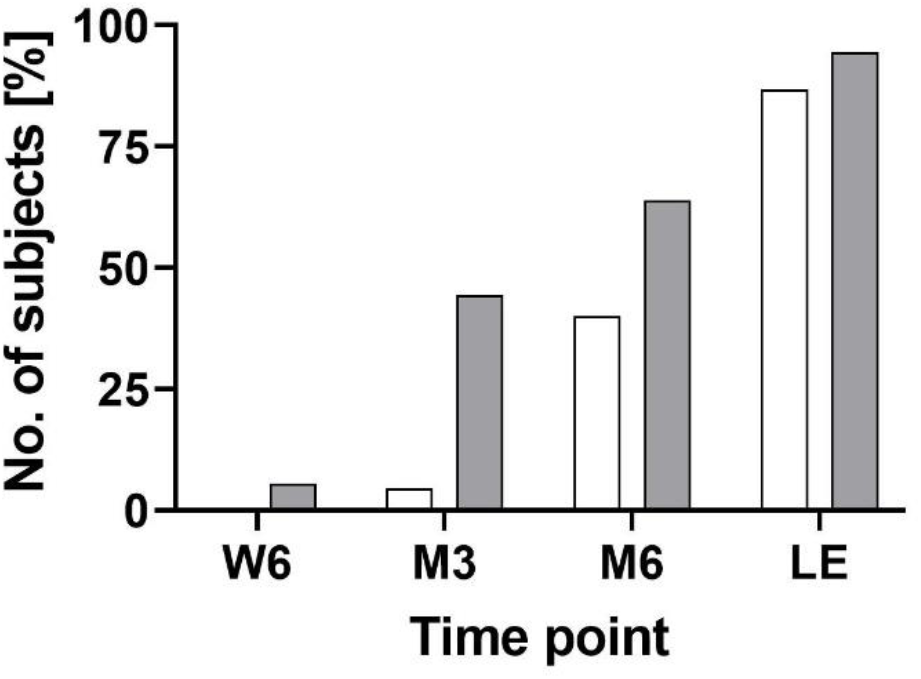
Relative number of subjects with unilateral, symptomatic, primary focal osteochondral lesions of the patella who were treated with respectively a combination of intra-articular injection of hyaluronic acid (HAi) and radial extracorporeal shock wave therapy (gray bars) or HAi alone (open bars) and individual improvement in VAS pain score by at least 60% at six weeks (W6), three months (M3) and six months (M6) after baseline, as well as at a last follow-up examination (LE) which was performed at 37.6 ± 1.7 (mean ± standard error of the mean) months after baseline (range, 12-59 months).

### Treatment success

Sixteen subjects (16/36 = 44.4%) who were treated with HAi+rESWT and two subjects (2/45 = 4.4%) who were treated with HAi alone showed individual improvement in the VAS pain score by at least 60% at M3. At W6, M6 and LE the numbers of subjects with treatment success were 2 (5.6%), 23 (63.9% and 34 (94.4%) in the cohort that was treated with HAi+rESWT, and 0 (0%), 18 (40%) and 39 (86.7%) in the cohort that was treated with HAi alone (Fig. 2). This difference between the cohorts was statistically significant (p = 0.007). Accordingly, the null hypothesis was rejected.

### WOMAC score

The mean WOMAC score of the subjects who were treated with HAi+rESWT decreased from 61.2 ± 1.7 at BL to 37.1 ± 0.9 at W6, 30.9 ± 0.7 at M3, 27.2 ± 0.8 at M6 and 20.5 ± 0.7 at LE, and of the subjects who were treated with HAi alone from 63.0 ± 1.1 at BL to 48.8 ± 1.2 at W6, 37.0 ± 1.1 at M3, 28.5 ± 0.8 at M6 and 23.1 ± 0.6 at LE (Fig. 1B). As in case of the VAS pain score the GLM-RM procedure showed a statistically significant interaction between Time and Treatment (Table 2). The Mann Whitney test showed statistically significant differences between the cohorts at W6, M3 and LE (Table 2).

### Area of the patella bone marrow edema on sagittal MRI scans

The mean PBME area of the subjects who were treated with HAi+rESWT decreased from 70.9 ± 2.8 mm^2^ at BL to 62.6 ± 2.7 mm^2^ at M3, 28.1 ± 2.3 mm^2^ at M6 and 18.0 ± 2.2 mm^2^ at LE, and of the subjects who were treated with HAi alone from 73.8 ± 1.5 mm^2^ at BL to 71.4 ± 1.4 mm^2^ at M3, 52.6 ± 1.2 mm^2^ at M6 and 31.7 ± 1.4 mm^2^ at LE (Fig. 1C). As in case of the VAS pain score and the WOMAC score the GLM-RM procedure showed a statistically significant interaction between Time and Treatment (Table 2). The Mann Whitney test showed statistically significant differences between the cohorts at M3, M6 and LE (Table 2). Three subjects who were treated with HAi+rESWT but no subject who was treated with HAi alone showed complete regression of PBME during the course of this study.

### Adverse events

Four subjects who were treated with HAi+rESWT (4/36=11.1%) reported mild complications such as transient reddening of the skin after a rESWT session, which disappeared within 24 hours after treatment. No serious adverse events were reported

## DISCUSSION

To our knowledge this is the first study on treatment of sPFOLP with a combination of HAi and rESWT. We evidenced the following key findings: (i) no severe adverse events related to the treatment of sPFOLP with HAi+rESWT in up to 59 months after treatment, (ii) significantly better outcome after treatment of sPFOLP with HAi+rESWT than after treatment with HAi alone, and (iii) treatment success documented by both subjective clinical endpoints (VAS pain score, WOMAC score) and an objective clinical endpoint (area of patella bone marrow edema on sagittal, fat suppression sequence MRI scans.

Symptomatic, primary focal osteochondral lesions of the patella are a manifestation of patellofemoral joint degeneration, and the conservative treatment of the disease mainly refers to the treatment principles of primary knee osteoarthrosis (knee OA). Both HAi and rESWT are established treatment modalities for knee OA [9-11, 23].

Hyaluronic acid is an intrinsic component of both the synovial fluid and the articular cartilage matrix. In osteoarthritis its unique rheological properties are lost because of a significant decrease in the molecular weight of HA, resulting in decreased viscosity of synovial fluid and disruption of cartilage [24]. A Cochrane systematic review published in 2006 found overall benefits to viscosupplementation (including but not limited to HA) in comparison to placebo for pain, function and patient global assessment scores [25, 26]. Another systematic review concluded that in patients with knee OA, viscosupplementation is associated with only small benefits that may be clinically irrelevant [27]. However, these authors demonstrated that high-molecular-weight and cross-linked preparations had greater effects than low-molecular-weight preparations [27]. Collectively, these data from the literature do not contradict our finding that HAi alone may not be the optimum treatment for sPFOLP.

Both focused ESWT (fESWT) and rESWT are used in treatment of knee OA [28]. Focused extracorporeal shock waves (fESWs) and rESWs are single acoustic impulses with an initial high positive peak pressure between 10 and 100 megapascals (MPa) reached in less than one microsecond [29, 30]. The positive pressure amplitude is followed by a low tensile amplitude of a few microseconds duration that can generate cavitation [30–32]. Further characteristics of extracorporeal shock waves are a short life cycle of approximately 10-20 µs and a broad frequency spectrum. Focused ESWs differ from rESWs in the penetration depth into the tissue, some physical characteristics, and the technique for generating them [15, 32, 33]. With respect to osteochondral lesions fESWs may be particularly useful for deep indications such as avascular necrosis of the hip, whereas rESWs may be more useful for superficial indications such as sPFOLP. Furthermore, a recent study demonstrated superiority of targeting subchondral bone over targeting articular cartilage in treatment of knee OA with ESWT [34]. Considering the anatomy of the patella and the fact that rESWs have their highest energy flux density (EFD) at the tip of the applicator [15, 31] (resulting in higher EFD in the subchondral bone than in the articular cartilage of the patella) rESWT appears more suitable for treating sPFOLP than fESWT. The well known negative correlation between increasing EFD of fESWs/rESWs and decreasing VAS pain score and decreasing WOMAC function index [23, 28, 35, 36] does not require to treat sPFOLP with fESWs (which can reach higher EFDs than rESWs) but to treat sPFOLP with adequate rESWT technology that can reach the necessary EFD (as applied in the present study as well as in [23] in the treatment of knee OA). Potential mechanisms of action of ESWT in knee OA are repeated mechanical stimulation of the lesion area [37], micro-injuries that may occur in the treated area [38] and stimulate bone remodeling [39, 40], and a number of chondroprotective effects including reduction in nitric oxide level and chondrocyte apoptosis, as well as increased VEGF, BMP-2 and osteocalcin in the subchondral bone [41–44]. As such, ESWT and HAi may have mutually beneficial effects in the treatment of sPFOLP. Whether ESWT can induce gene expression of HA (as demonstrated for lubricin [45], another molecule that serves as lubricant in synovial fluid and is decreased in OA [46]) is currently unknown.

Our finding of substantial reduction of patella bone marrow edema after treating sPFOLP with HAi+rESWT (on average −60% at M6 and −75% at the time of the last follow-up examination) is in line with a recent report of reduction of bone marrow edema in knee OA after treatment with fESWT [47]. However, in the latter study this effect was achieved using fESWs at very high EFD (>0.44 mJ/mm2) which required surface anesthesia (lidocaine hydrochloride gel) and intravenous application of flurbiprofen axetil. This is in contrast to the present study in which rESWT could be administered without anesthesia or any other analgesic.

## Limitations

The present study has a number of limitations. Specifically, we performed a retrospective two-cohort study rather than a prospective RCT. As a result, subjects were not randomly allocated to either treatment. Furthermore, subjects, therapists and outcome assessors were not blinded because this study relied on the analysis of completed medical records. Thus, our findings should be verified in adequate RCTs. Another limitation is the fact that no subjects were treated with rESWT alone. However, a recent RCT demonstrated that treatment of knee OA with respectively fESWT (3 sessions, one session per week, 1000 fESWs per session with EFD of 0.05 mJ/mm^2^) or HAi (3 injections, one injection per week, 20 mg sodium hyaluronate in 2 ml liquid) resulted in similar improvement of mean VAS pain score and mean WOMAC score at one and three months after baseline [48] (of note, the results of [48] and the present study cannot be directly compared with each other because in [48] the mean VAS score and the mean WOMAC score were lower than in the present study). Considering the different mechanisms of HA and ESWT in OA it appears unlikely that rESWT alone would cause the same clinical benefits as HAi+rESWT as shown in the present study.

## Conclusions

The results of this study suggest that the use of HAi+rESWT in subjects with sPFOLP is safe and leads to better outcome than HAi alone, without adverse effects. Adequate RCTs are necessary to verify this. Clinicians should consider HAi+rESWT instead of HAi alone in the management of sPFOLP.

## Data Availability

The datasets used and analyzed during this study are available from the corresponding author on reasonable request, taking into account any confidentiality.

## Abbreviations

BL: baseline
BMI: body mass index
EFD: energy flux density
EFD+: positive energy flux density
ESWT: extracorporeal shock wave therapy
F: Fisher’s exact test
fESWs: focused extracorporeal shock waves
fESWT: focused extracorporeal shock wave therapy
GLM-RM: general linear model repeated measures
HA: hyaluronic acid
HAi: intra-articular injection of hyaluronic acid
kg: kilogram
LE: last follow-up examination
M3: three months after baseline
m^2^: squared meter
M6: six months after baseline
max: maximum value
min: minimum value
mJ/mm2: millijoule per squared millimeter
ml: milliliter
MRI: magnetic resonance imaging
OA: osteoarthritis
PBME: patellar bone marrow edema
rESWs: radial extracorporeal shock waves
rESWT: radial extracorporeal shock wave therapy
RCT: randomized controlled trial
SD: standard deviation
SEM: standard error of the mean
sPFOLP: symptomatic, primary focal osteochondral lesions of the patella
T: unpaired t test
VAS: visual analog scale
W6: six weeks
WOMAC: Western Ontario and McMaster Universities Osteoarthritis Index

## Acknowledgements

We express our gratitude to all the subjects with SPFOLP who were treated at the outpatient department of our hospital between January 1^st^, 2014 and January 31^th^, 2018 and whose data were considered in this retrospective study. We also thank everyone at the Center for Joint Surgery, Southwest Hospital, Third Military Medical University (Army Medical University), Chongqing, China for their contribution to the study operations.

## Funding

This study did not receive any external funding.

## Authors’ contributions

JC, CZ, LY and XD conceived this retrospective study and participated in the design of this retrospective study. HH performed the treatments. JC and CZ analyzed the MRI scans. CS performed the statistical analysis and drafted the manuscript. JC and XD helped to draft the manuscript. All authors read and approved the final manuscript.

## Ethics approval and consent to participate

Ethical approval for this retrospective study was granted by the Ethics Committee of the Southwest Hospital, Third Military Medical University (Army Medical University), Chongqing, China (registration number KY201915). Because the study design did not require any additional interaction with the subjects there was no need to obtain consent for participation in this retrospective study according to the rules and regulations of Southwest Hospital, Third Military Medical University (Army Medical University), Chongqing 400038, China.

## Consent for publication

Because no any data and images are shown that could be used to identify individual subjects in this study there was no need to obtain consent for publication.

## Competing interests

This study was performed with the radial extracorporeal shock wave device Swiss DolorClast, which is manufactured and distributed by Electro Medical Systems (Nyon, Switzerland). CS has received research funding at LMU Munich and consulted (until December 31^st^, 2017) for Electro Medical Systems. However, Electro Medical Systems had no role in study design, data collection, data analysis, data interpretation, or writing of the report. All other authors declare no competing interests.

## References

1. Buda R, Vannini F, Cavallo M, Grigolo B, Cenacchi A, Giannini S. Osteochondral lesions of the knee: a new one-step repair technique with bone-marrow-derived cells. J Bone Joint Surg Am. 2010;92 Suppl 2:2–11. doi: 10.2106/JBJS.J.00813.

2. Figueroa D, Meleán P, Calvo R, Gili F, Zilleruelo N, Vaisman A. Osteochondral autografts in full thickness patella cartilage lesions. Knee. 2011;18(4):220–223. doi: 10.1016/j.knee.2010.05.016.

3. Kao YJ, Ho J, Allen CR. Evaluation and management of osteochondral lesions of the knee. Phys Sportsmed. 2011;39(4):60–69. doi: 10.3810/psm.2011.11.1940.

4. Dall’Oca C, Cengarle M, Costanzo A, Giannini N, Vacchiano A, Magnan B. Current concepts in treatment of early knee osteoarthritis and osteochondral lesions; the role of biological augmentations. Acta Biomed. 2017;88(4S):5–10. doi: 10.23750/abm.v88i4-S.6788.

5. Cotter EJ, Hannon CP, Christian DR, Wang KC, Lansdown DA, Waterman BR, Frank RM, Cole BJ. Clinical outcomes of multifocal osteochondral allograft transplantation of the knee: an analysis of overlapping grafts and multifocal lesions. Am J Sports Med. 2018;46(12):2884–2893. doi: 10.1177/0363546518793405.

6. Degen RM, Coleman NW, Tetreault D, Chang B, Mahony GT, Camp CL, Anthony SG, Williams RJ. Outcomes of patellofemoral osteochondral lesions treated with structural grafts in patients older than 40 years. Cartilage. 2017;8(3):255–262. doi: 10.1177/1947603516665441.

7. Grelsamer RP, Dejour D, Gould J. The pathophysiology of patellofemoral arthritis. Orthop Clin North Am. 2008;39(3):269–274. doi: 10.1016/j.ocl.2008.03.001.

8. Gorbachova T, Melenevsky Y, Cohen M, Cerniglia BW. Osteochondral lesions of the knee: differentiating the most common entities at MRI. Radiographics. 2018;38(5):1478–1495. doi: 10.1148/rg.2018180044.

9. Sherman SL, Thomas DM, Farr J. Chondral and osteochondral lesions in the patellofemoral joint: when and how to manage. Ann Joint 2018;3:53. doi: 10.21037/aoj.2018.04.12.

10. Evanich JD, Evanich CJ, Wright MB, Rydlewicz JA. Efficacy of intraarticular hyaluronic acid injections in knee osteoarthritis. Clin Orthop Relat Res. 2001;(390):173–181. doi: 10.1097/00003086-200109000-00020.

11. Kon E, Mandelbaum B, Buda R, Filardo G, Delcogliano M, Timoncini A, Fornasari PM, Giannini S, Marcacci M. Platelet-rich plasma intra-articular injection versus hyaluronic acid viscosupplementation as treatments for cartilage pathology: from early degeneration to osteoarthritis. Arthroscopy. 2011;27(11):1490–1501. doi: 10.1016/j.arthro.2011.05.011.

12. Chahla J, Hinckel BB, Yanke AB, Farr J; Metrics of Osteochondral Allografts (MOCA) Group, Bugbee WD, Carey JL, Cole BJ, Crawford DC, Fleischli JE, Getgood A, Gomoll AH, Gortz S, Gross AE, Jones DG, Krych AJ, Lattermann C, Mandelbaum BR, Mandt PR, Minas T, Mirzayan R, Mologne TS, Polousky JD, Provencher MT, Rodeo SA, Safir O, Sherman SL, Strauss ED, Strickland SM, Wahl CJ, Williams RJ 3rd. An expert consensus statement on the management of large chondral and osteochondral defects in the patellofemoral joint. Orthop J Sports Med. 2020;8(3):2325967120907343. doi: 10.1177/2325967120907343.

13. Hinckel BB, Pratte EL, Baumann CA, Gowd AK, Farr J, Liu JN, Yanke AB, Chahla J, Sherman SL. Patellofemoral cartilage restoration: a systematic review and meta-analysis of clinical outcomes. Am J Sports Med. 2020;48(7):1756–1772. doi: 10.1177/0363546519886853.

14. Speed C. A systematic review of shockwave therapies in soft tissue conditions: focusing on the evidence. Br J Sports Med. 2014;48(21):1538–1542. doi: 10.1136/bjsports-2012-091961.

15. Schmitz C, Császár NB, Milz S, Schieker M, Maffulli N, Rompe JD, Furia JP. Efficacy and safety of extracorporeal shock wave therapy for orthopedic conditions: a systematic review on studies listed in the PEDro database. Br Med Bull. 2015;116(1):115–138. doi: 10.1093/bmb/ldv047.

16. Reilly JM, Bluman E, Tenforde AS. Effect of shockwave treatment for management of upper and lower extremity musculoskeletal conditions: a narrative review. PM R. 2018;10(12):1385–1403. doi: 10.1016/j.pmrj.2018.05.007.

17. Wang CJ, Wang FS, Huang CC, Yang KD, Weng LH, Huang HY. Treatment for osteonecrosis of the femoral head: comparison of extracorporeal shock waves with core decompression and bone-grafting. J Bone Joint Surg Am. 2005;87(11):2380–2387. doi: 10.2106/JBJS.E.00174.

18. Lee JY, Kwon JW, Park JS, Han K, Shin WJ, Lee JG, Lee BH. Osteonecrosis of femoral head treated with extracorporeal shock wave therapy: analysis of short-term clinical outcomes of treatment with radiologic staging. Hip Pelvis. 2015 Dec;27(4):250–7. doi: 10.5371/hp.2015.27.4.250. Epub 2015 Dec 30. PMID: 27536633; PMCID: PMC4972796.

19. Algarni AD, Al Moallem HM. Clinical and radiological outcomes of extracorporeal shock wave therapy in early-stage femoral head osteonecrosis. Adv Orthop. 2018;2018:7410246. doi: 10.1155/2018/7410246.

20. Wang QW, Zhang QY, Gao FQ, Sun W. Focused extra-corporeal shockwave treatment during early stage of osteonecrosis of femoral head. Chin Med J. 2019;132(15):1867–1869. doi: 10.1097/CM9.0000000000000331.

21. Zhang C, Huang H, Yang L, Duan X. Extracorporeal shock wave therapy for pain relief after arthroscopic treatment of osteochondral lesions of talus. J Foot Ankle Surg. 2020;59(1):190–194. doi: 10.1053/j.jfas.2019.07.015.

22. Moher D, Hopewell S, Schulz KF, Montori V, Gøtzsche PC, Devereaux PJ, Elbourne D, Egger M, Altman DG; CONSORT. CONSORT 2010 explanation and elaboration: updated guidelines for reporting parallel group randomised trials. Int J Surg. 2012;10(1):28–55. doi: 10.1016/j.ijsu.2011.10.001.

23. Zhao Z, Jing R, Shi Z, Zhao B, Ai Q, Xing G. Efficacy of extracorporeal shockwave therapy for knee osteoarthritis: a randomized controlled trial. J Surg Res. 2013;185(2):661–666. doi: 10.1016/j.jss.2013.07.004.

24. Abatangelo G, Vindigni V, Avruscio G, Pandis L, Brun P. Hyaluronic acid: redefining its role. Cells. 2020;9(7):E1743. doi: 10.3390/cells9071743.

25. Bellamy N, Campbell J, Robinson V, Gee T, Bourne R, Wells G. Viscosupplementation for the treatment of osteoarthritis of the knee. Cochrane Database Syst Rev. 2006;(2):CD005321. doi: 10.1002/14651858.CD005321.pub2.

26. Evaniew N, Simunovic N, Karlsson J. Cochrane in CORR^®^: viscosupplementation for the treatment of osteoarthritis of the knee. Clin Orthop Relat Res. 2014;472(7):2028–2034. doi: 10.1007/s11999-013-3378-8.

27. Rutjes AW, Jüni P, da Costa BR, Trelle S, Nüesch E, Reichenbach S. Viscosupplementation for osteoarthritis of the knee: a systematic review and meta-analysis. Ann Intern Med. 2012;157(3):180–191. doi: 10.7326/0003-4819-157-3-201208070-00473.

28. Liao CD, Tsauo JY, Liou TH, Chen HC, Huang SW. Clinical efficacy of extracorporeal shockwave therapy for knee osteoarthritis: a systematic review and meta-regression of randomized controlled trials. Clin Rehabil. 2019;33(9):1419–1430. doi: 10.1177/0269215519846942.

29. Rompe JD, Furia J, Weil L, Maffulli N. Shock wave therapy for chronic plantar fasciopathy. Br Med Bull. 2007;81-82:183–208. doi: 10.1093/bmb/ldm005.

30. Hochstrasser T, Frank HG, Schmitz C. Dose-dependent and cell type-specific cell death and proliferation following in vitro exposure to radial extracorporeal shock waves. Sci Rep. 2016;6:30637. doi: 10.1038/srep30637.

31. Császár NB, Angstman NB, Milz S, Sprecher CM, Kobel P, Farhat M, Furia JP, Schmitz C. Radial shock wave devices generate cavitation. PLoS One. 2015;10(10):e0140541. doi: 10.1371/journal.pone.0140541.

32. Schmitz C, Császár NB, Rompe JD, Chaves H, Furia JP. Treatment of chronic plantar fasciopathy with extracorporeal shock waves (review). J Orthop Surg Res. 2013;8:31. doi:10.1186/1749-799X-8-31.

33. Ogden JA, Tóth-Kischkat A, Schultheiss R. Principles of shock wave therapy. Clin Orthop Relat Res. 2001;(387):8–17. doi: 10.1097/00003086-200106000-00003.

34. Chou WY, Cheng JH, Wang CJ, Hsu SL, Chen JH, Huang CY. Shockwave targeting on subchondral bone is more suitable than articular cartilage for knee osteoarthritis. Int J Med Sci. 2019;16(1):156–166. doi: 10.7150/ijms.26659.

35. Hammam RF, Kamel RM, Draz AH, Azzam AA, Abu El Kasem ST. Comparison of the effects between low-versus medium-energy radial extracorporeal shock wave therapy on knee osteoarthritis: A randomised controlled trial. J Taibah Univ Med Sci. 2020;15(3):190–196. doi: 10.1016/j.jtumed.2020.04.003.

36. Imamura M, Alamino S, Hsing WT, Alfieri FM, Schmitz C, Battistella LR. Radial extracorporeal shock wave therapy for disabling pain due to severe primary knee osteoarthritis. J Rehabil Med. 2017;49(1):54–62. doi: 10.2340/16501977-2148.

37. Huang C, Holfeld J, Schaden W, Orgill D, Ogawa R. Mechanotherapy: revisiting physical therapy and recruiting mechanobiology for a new era in medicine. Trends Mol Med. 2013;19(9):555–564. doi: 10.1016/j.molmed.2013.05.005.

38. Da Costa Gómez TM, Radtke CL, Kalscheur VL, Swain CA, Scollay MC, Edwards RB, Santschi EM, Markel MD, Muir P. Effect of focused and radial extracorporeal shock wave therapy on equine bone microdamage. Vet Surg. 2004;33(1):49–55. doi: 10.1111/j.1532-950x.2004.040005.x.

39. Burr DB. Targeted and nontargeted remodeling. Bone. 2002;30(1):2–4. doi: 10.1016/s8756-3282(01)00619-6.

40. Wang CJ, Weng LH, Ko JY, Sun YC, Yang YJ, Wang FS. Extracorporeal shockwave therapy shows chondroprotective effects in osteoarthritic rat knee. Arch Orthop Trauma Surg. 2011;131(8):1153–1158. doi: 10.1007/s00402-011-1289-2.

41. Ma HZ, Zeng BF, Li XL. Upregulation of VEGF in subchondral bone of necrotic femoral heads in rabbits with use of extracorporeal shock waves. Calcif Tissue Int. 2007;81(2):124–131. doi: 10.1007/s00223-007-9046-9.

42. Wang CJ, Hsu SL, Weng LH, Sun YC, Wang FS. Extracorporeal shockwave therapy shows a number of treatment related chondroprotective effect in osteoarthritis of the knee in rats. BMC Musculoskelet Disord. 2013;14:44. doi: 10.1186/1471-2474-14-44.

43. Zhao Z, Ji H, Jing R, Liu C, Wang M, Zhai L, Bai X, Xing G. Extracorporeal shock-wave therapy reduces progression of knee osteoarthritis in rabbits by reducing nitric oxide level and chondrocyte apoptosis. Arch Orthop Trauma Surg. 2012;132(11):1547–1553. doi: 10.1007/s00402-012-1586-4.

44. Wang CJ, Sun YC, Wong T, Hsu SL, Chou WY, Chang HW. Extracorporeal shockwave therapy shows time-dependent chondroprotective effects in osteoarthritis of the knee in rats. J Surg Res. 2012;178(1):196–205. doi: 10.1016/j.jss.2012.01.010.

45. Zhang D, Kearney CJ, Cheriyan T, Schmid TM, Spector M. Extracorporeal shockwave-induced expression of lubricin in tendons and septa. Cell Tissue Res. 2011;346(2):255–262. doi: 10.1007/s00441-011-1258-7.

46. Kosinska MK, Ludwig TE, Liebisch G, Zhang R, Siebert HC, Wilhelm J, Kaesser U, Dettmeyer RB, Klein H, Ishaque B, Rickert M, Schmitz G, Schmidt TA, Steinmeyer J. Articular joint lubricants during osteoarthritis and rheumatoid arthritis display altered levels and molecular species. PLoS One. 2015;10(5):e0125192. doi: 10.1371/journal.pone.0125192.

47. Kang S, Gao F, Han J, Mao T, Sun W, Wang B, Guo W, Cheng L, Li Z. Extracorporeal shock wave treatment can normalize painful bone marrow edema in knee osteoarthritis: A comparative historical cohort study. Medicine. 2018;97(5):e9796. doi: 10.1097/MD.0000000000009796.

48. Lee JK, Lee BY, Shin WY, An MJ, Jung KI, Yoon SR. Effect of extracorporeal shockwave therapy versus intra-articular injections of hyaluronic acid for the treatment of knee osteoarthritis. Ann Rehabil Med. 2017;41(5):828–835. doi: 10.5535/arm.2017.41.5.828.

